# Adverse childhood experiences across Multiracial and monoracial groups with and without Indigenous ancestry

**DOI:** 10.1101/2023.09.25.23296116

**Authors:** Tracy Lam-Hine, Corinne A. Riddell, Patrick T. Bradshaw, Michael Omi, Amani M. Allen

## Abstract

Adverse childhood experiences (ACEs) are linked to increased risk of a host of health outcomes in adulthood. Descriptive ACEs prevalence studies have previously identified that Multiracial people have the highest mean ACE score of any racial group, but do not offer explanations for these disparities. However, Multiracial people form one of the fastest-growing populations in the US, and the largest subgroup of Multiracial people is those that claim American Indian/Native American (AI/NA) ancestry. Mean ACE counts (scores) are also high among the AI/NA population, which may reflect impacts of land occupation and structural racism. This descriptive study uses data from the National Longitudinal Study of Adolescent to Adult Health (Add Health) to test the hypothesis that mean ACE scores and prevalence of ACE components are higher among Multiracial AI/NA participants than Multiracial non-AI/NA participants. Mean scores were highest among AI/NA (mean = 3.21, 95% CI: 2.54, 3.97), Multiracial AI/NA (2.95, 95% CI: 2.71, 3.18), Multiracial non-AI/NA (2.88, 95% CI: 2.57, 3.19), and Black (2.84, 95% CI: 2.65, 3.02) groups. Differences in mean ACE scores and prevalence of ACE components between the two Multiracial groups were all insignificant. Results from this study did not support our hypothesis, suggesting that the Multiracial population’s high ACE scores are not driven primarily by those with AI/NA ancestry. Future studies should repeat this analysis in larger datasets and explore other determinants of high mean ACE scores among the Multiracial population.

## Background

Adverse childhood experiences (ACEs) are traumatic events occurring during childhood known to be associated with the development of physical, psychological, and behavioral health problems later in life.^1,2^ Strong dose-response effects of ACEs across multiple domains of health outcomes have been documented. One systematic review and meta-analysis found those reporting four or more ACEs were at higher risk for every studied health condition compared to those reporting no ACEs, with associations strongest for psychological and behavioral health outcomes and weaker for cancer and cardiometabolic outcomes.^1^ Clinical guidance for ACEs risk assessments suggest that the risk of poor health tends to increase sharply at a score of three or four ACEs, depending on the outcome.^3^ However, others have argued that ACE scores are more appropriately used to assess population-level differences in health risks, rather than prediction of a specific individual’s future health.^4^

The initial ACEs study conducted by Kaiser Permanente between 1995-97 found widespread prevalence of ACEs in a relatively high socioeconomic status (SES), commercially-insured population in San Diego, CA.^5^ The original ten ACE domains include: emotional, physical, and sexual abuse, emotional and physical neglect, parental separation or divorce, mother treated violently, household substance abuse, mental illness in household, and incarceration of a parent. The study substantiated the need for wider screening for traumatic childhood events beyond low SES populations and communities of color. However, disparities in the prevalence of ACEs by race, SES, sexual orientation, and other characteristics persist, with populations in socially-privileged positions consistently reporting fewer ACEs than marginalized groups.^6^

Discussions on the causes of racial disparities in ACEs are limited, which may reflect the reality that ACEs are most frequently used as an individual screening tool in medicine rather than as a measure of population health or health equity.^7^ Additionally, ACEs are not a discrete condition, but a disparate set of traumatic events, which may also explain the paucity of literature on causes. The few studies that discuss racial disparities in ACEs suggest institutional and/or structural or systemic racism as potential causes.^8–10^ Racism becomes institutionalized when social institutions — such as schools, employers, courts, etc. — adopt and enforce policies or practices that are either overtly racist or lead to racial disparities in outcomes. Some examples include: state-sponsored occupation of Indigenous land, racially discriminatory lending practices, overpolicing and mass incarceration of people of color, and racialized treatment standards in healthcare delivery. Structural or systemic racism reflects the cumulative effects of racism across interacting institutions, enabling preservation of White privilege across space and time, and is considered a fundamental cause of health inequities.^11–14^ Some forms of structural and institutional racism cause ACEs directly; for example, overpolicing and mass incarceration in Black and Brown communities results in large numbers of children of color with parents in prison.^15^ Other forms may cause ACEs indirectly via material deprivation and the stress pathway across time. For example, the colonization and occupation of Indigenous lands and the subsequent forced resettlement of American Indian/Alaska Native (AI/NA) peoples into reservations with substandard educational and employment opportunities has impacted generations of AI/NA families. The stress on AI/NA families resulting from these structurally racist events predisposes their children to adverse developmental environments and experiences.^16,17^

Multiple studies have found that Multiracial — those identifying with more than one racial group — and AI/NA people consistently report the highest number of ACEs of all race/ethnicities.^6,9,18–21^ However, small population sizes and inconsistency of racial data collection instruments mean that such population health analyses infrequently focus on Multiracial and AI/NA groups. As described above, there is a clear connection between colonialist land occupation, structural racism, and ACEs for the AI/NA population. However, most literature on structural racism discusses its impact on monoracial populations of color, making it unclear if structural racism is also responsible for elevated ACEs among the Multiracial populations.^22^

None of the studies identifying disparities in ACEs between monoracial and Multiracial populations discuss possible reasons for this difference. Given high rates of ACEs also reported in the AI/NA population and the disproportionate effects of structural racism and land occupation burdening this community, the Multiracial-monoracial disparity could be a result of the high numbers of Multiracial people claiming AI/NA ancestry. Disaggregation of Multiracial ancestry to investigate this theory is not possible in publicly-available versions of large, nationally representative public health datasets, such as the National Health Interview Survey (NHIS), the Behavioral Risk Factor Surveillance System (BRFSS), and the National Survey of Children’s Health (NSCH), as the race variables provided collapse detailed multiple race responses into a single “Multiracial” category. Investigating disparities in ACEs in datasets that allow for detailed disaggregation of the Multiracial category could be a first step in better understanding the unique and urgent health risks facing this population.

To examine factors potentially associated with the disparity in exposure to ACEs between Multiracial and monoracial groups, in this study we test the hypothesis that Multiracial participants with AI/NA ancestry will report significantly higher mean ACE score and prevalence of ACE components than Multiracial participants without AI/NA ancestry. Support of this hypothesis would suggest that the Multiracial population’s high mean ACE score is driven by the large proportion of those with AI/NA ancestry, possibly reflecting the multigenerational consequences of colonialist land occupation affecting AI/NA communities.

## Methods

### Data and analytic sample

The National Longitudinal Study of Adolescent to Adult Health (Add Health) is a longitudinal, nationally representative study following over 20,000 individuals from grades 7-12 in 1994-95 through four waves of follow-up (1996, 2001-02, 2008-09, 2016-18).^23^ Add Health is the largest nationally representative longitudinal study that allows study participants to select more than one racial category, making it an important source of research on Multiracial people.^24,25^ Participants were recruited through a school-based cluster sample, sampled with unequal probability and without stratification. From a database of all 26,666 high schools in the US, eighty public and private high schools and associated feeder middle schools were selected for size, type, grade range, setting, demographics, and geographic location. Within schools, specific racial/ethnic groups and students with disabilities were oversampled for population weighting. Students in grades 7-12 were sampled from school enrollment rosters and invited to complete an at-home interview during Wave 1, with a total sample of 20,745 participants. Wave 1 included detailed questions about the adolescent’s demographics and family background, social networks, home and school environments, and health behaviors. Wave 4 was conducted 14 years after Wave 1 in 2008-09, when the participants were in their late 20’s and included measurements of the participants’ metabolic and cardiovascular function. More details about the study design can be found in the Add Health study documentation.^26^

### Classification of race

Add Health participants were asked to self-identify their race and ethnicity in Waves 1 and 3. In Wave 1, participants were given five choices of racial categories: White, Black, American Indian/Native American (here reclassified as AI/NA), Asian, and Other, and were given the option to select multiple categories. In a separate question, participants were asked to indicate if they identified as Hispanic or Latino. At Wave 3, the “Other” option was removed, leaving respondents to select from four race options: White, Black, AI/NA, and Asian, still with the option to select multiple categories. Given the conceptual and methodological complexities of enumerating Multiracial people in quantitative data, we aimed to maximize the number of participants who classified as Multiracial. If a participant identified as Multiracial in both waves, we used their Wave 3 race. If a participant only identified as Multiracial in Wave 1 (but not in Wave 3), we used their Wave 1 race. If a Multiracial participant selected “Other” in Wave 1, we used their Wave 1 racial combination.

Otherwise, we used their Wave 3 race. We further split the Multiracial group into those reporting and not reporting AI/NA ancestry. The final racial categories were White, Black, Asian, AI/NA, Multiracial AI/NA, and Multiracial non-AI/NA. Post-hoc analysis led us to exclude the “Other alone” category due to the much smaller sample size relative to other groups (n = 24) and large confidence intervals, preventing meaningful interpretation of results.

Despite official attempts to separate race and Hispanic ethnicity, a majority of Hispanic/Latino people consider Latinidad to be a core part of their racial identity.^27^ When Hispanic ethnicity is assessed separately from race, it becomes impossible to differentiate between monoracial or Multiracial Hispanic/Latino people. Given these challenges, we align our methods with other studies^28^ using Add Health data that have excluded participants that identified as Hispanic/Latino in Wave 1 or Wave 3.

### Measurement of ACEs

The original ACEs questionnaire included ten questions each covering a single domain of adverse experiences^29^: (1) emotional abuse, (2) physical abuse, (3) sexual abuse, (4) emotional neglect, (5) physical neglect, (6) parental separation or divorce, (7) mother treated violently, (8) household substance abuse, (9) mental illness in household, and (10) incarceration of household member. The original ACEs questionnaire was not administered in any Add Health surveys. Instead, researchers have used Add Health questions that approximate those asked in the original ACEs questionnaire.^30–32^ We used a modified version of the widest set of questions available in Add Health data to construct ten variables approximating the components of the original ACE score, following the methods of Lee et al. and Otero.^33,34^ We coded variables as binary and then summed them, with a minimum of zero and maximum of ten. Further details on the questions and variables used to construct the ACEs score in Add Health are available in Appendix A.

### Statistical analyses

We compared mean ACE scores (e.g.: the mean number of ACEs experienced) and unadjusted prevalence ratios of reporting each ACE component across each racial group. Because the Add Health design specifically oversampled certain groups based on demographic characteristics, we conducted a design-based analysis to address selection bias built into the sampling plan. We used complex survey weights to produce nationally-representative estimates and standard errors. We assessed differences in group mean scores and prevalence of individual ACE components using Tukey’s honest significance test and by comparing 95% confidence intervals around mean score estimates.^35,36^ We adjusted p-values using Bonferroni correction for multiple testing. All data and statistical analyses were performed in R using the “tidyverse” and “multcomp” packages.^37–40^ Analysis of design effects with complex survey weights was implemented using the “survey” package.^41^ There were high levels of missingness in ACE components (frequencies presented in Appendix B); we thus performed Markov chain multiple imputation with 30 iterations and 4 chains using the “mi” and “mitools” packages.^41,42^ Our imputation models specified all ACE components along with participant demographic information and hypothesized missingness predictor variables; results were pooled across 20 imputed datasets.

## Results

Unweighted counts and weighted joint distributions of participant sex, age, component ACE scores, and summary ACE scores by racial group are presented in Table 1.

**Table 1.** Participant characteristics and ACE components^a^ stratified by race, Add Health 1994-2008.

| **Characteristic** | **Overall** | **White** | **Black** | **Asian** | **AI/NA** | **M. AI/NA** | **M. non-AI/NA** |
| --- | --- | --- | --- | --- | --- | --- | --- |
|  | 12,372 (100%) | 7,742 (74%) | 2,915 (17%) | 805 (3.2%) | 76 (0.6%) | 484 (3.9%) | 313 (2.5%) |
| Male sex | 5,778 (51%) | 3,672 (51%) | 1,267 (50%) | 418 (53%) | 41 (64%) | 230 (50%) | 150 (50%) |
| Age | 29.0 | 28.9 | 29.2 | 29.2 | 28.8 | 28.8 | 28.8 |
| Emotional abuse | 5,779 (47%) | 3,571 (45%) | 1,329 (51%) | 406 (51%) | 33 (45%) | 268 (52%) | 172 (54%) |
| Physical abuse | 2,825 (28%) | 1,674 (26%) | 603 (28%) | 264 (37%) | 22 (36%) | 164 (37%) | 98 (37%) |
| Sexual abuse | 461 (5.9%) | 246 (5.2%) | 133 (8.2%) | 37 (5.8%) | 4 (8.4%) | 34 (9.4%) | 7 (4.6%) |
| Emotional neglect | 3,715 (30%) | 2,209 (28%) | 903 (32%) | 281 (35%) | 27 (36%) | 186 (39%) | 109 (33%) |
| Physical neglect | 4,097 (43%) | 2,453 (42%) | 954 (47%) | 330 (53%) | 29 (46%) | 212 (48%) | 119 (47%) |
| Parental divorce or separation | 3,019 (30%) | 1,922 (30%) | 774 (34%) | 64 (14%) | 17 (34%) | 149 (31%) | 93 (35%) |
| Mother treated violently | 231 (6.4%) | 148 (5.2%) | 61 (12.4%) | 6 (3.0%) | 0 (3.6%) | 6 (3.6%) | 10 (12%) |
| Household substance abuse | 1,949 (22%) | 1,302 (21%) | 408 (23%) | 49 (13%) | 23 (48%) | 118 (32%) | 49 (26%) |
| Household mental illness | 1,187 (16%) | 706 (14%) | 313 (24%) | 70 (14%) | 14 (31%) | 54 (18%) | 30 (14%) |
| Parental incarceration | 2,206 (19%) | 1,212 (16%) | 726 (29%) | 48 (6.3%) | 24 (32%) | 124 (25%) | 72 (26%) |
| Mean ACE score | 2.46 | 2.35 | 2.84 | 2.32 | 3.21 | 2.95 | 2.84 |

Abbreviations: ACE = adverse childhood experience; M. = Multiracial; AI/NA = American Indian/Native American
<sup>a</sup> Counts are crude non-missing values; proportions (for categorical variables) and means (for continuous) are pooled estimates from 20 survey-weighted imputations

Total sample size was 12,372 after removing Hispanic/Latino participants, “Other alone” participants, and those missing complex survey weights for design-based analysis. Of these, 9,538 (94.2%) were monoracial and 834 (5.82%) were Multiracial. Monoracial White (n = 7,742, 74%) and Black (n = 2,915, 17%) participants form the largest groups in the study sample. Among those identifying as Multiracial, 484 (61%) indicated that they had some AI/NA ancestry, while 313 (29%) did not.

Despite not having the highest mean ACE score, non-AI/NA Multiracial participants had the highest prevalence of four different ACE components – more than any other group; 54% reported emotional abuse, 37% physical abuse (equal to Asian and Multiracial AI/NA participants), 35% parental divorce or separation, and 12% mother treated violently (equal to Black participants). AI/NA and Multiracial AI/NA participants reported the highest prevalence each of three different components (AI/NA: 48% household substance, 31% household mental illness, 32% parental incarceration; Multiracial AI/NA: 37% physical abuse, 9.4% sexual abuse, 39% emotional neglect). Asian participants had the highest prevalence of physical neglect (53%) and similarly high levels of physical abuse to the Multiracial groups (37%). A similarly high proportion of Black participants reported their mothers being treated violently to non-AI/NA Multiracial participants (12%).

Among all racial groups, AI/NA (mean = 3.21, 95% CI: 2.54, 3.97), Multiracial AI/NA (2.95, 95% CI: 2.71, 3.18), Multiracial non-AI/NA (2.88, 95% CI: 2.57, 3.19), and Black (2.84, 95% CI: 2.65, 3.02) participants reported the highest mean ACE scores. Formal pairwise tests of difference in mean ACE score between racial groups are presented in Table 2. Difference in mean scores were not significant between the two Multiracial groups, nor in any pairwise comparisons between these groups and the AI/NA or Black groups. The mean scores of these four groups were all significantly higher than those of the White or Asian groups.

**Table 2.** Summary of group mean ACE scores and Tukey’s honest significance pairwise tests of group difference in mean ACE scores, Add Health 1994-2008.

| Reference | Mean ACE score (95% CI) | Comparison | Mean Diff (95% CI) | p-value |
| --- | --- | --- | --- | --- |
| White | 2.35 (2.26, 2.44) | Black | 0.33 (0.25, 0.42) | < 0.001 |
|  |  | Asian | -0.07 (-0.21, 0.07) | 0.325 |
|  |  | AI/NA | 0.43 (0.22, 0.64) | < 0.001 |
|  |  | M. AI/NA | 0.62 (0.46, 0.78) | < 0.001 |
|  |  | M. non-AI/NA | 0.53 (0.33, 0.73) | < 0.001 |
| Black | 2.84 (2.65, 3.02) | Asian | -0.40 (-0.56, -0.25) | < 0.001 |
|  |  | AI/NA | 0.10 (-0.12, 0.32) | 0.390 |
|  |  | M. AI/NA | 0.29 (0.11, 0.46) | 0.001 |
|  |  | M. non-AI/NA | 0.18 (-0.01, 0.40) | 0.062 |
| Asian | 2.32 (2.09, 2.54) | AI/NA | 0.50 (0.25, 0.76) | < 0.001 |
|  |  | M. AI/NA | 0.69 (0.49, 0.89) | < 0.001 |
|  |  | M. non-AI/NA | 0.60 (0.36, 0.84) | < 0.001 |
| AI/NA | 3.21 (2.54, 3.97) | M. AI/NA | 0.19 (-0.08, 0.46) | 0.164 |
|  |  | M. non-AI/NA | -0.10 (-0.18, 0.38) | 0.486 |
| M. AI/NA | 2.95 (2.71, 3.18) | M. non-AI/NA | -0.09 (-0.33, 0.15) | 0.470 |
| M. non-AI/NA | 2.88 (2.57, 3.19) |  |  |  |

Abbreviations: M. = Multiracial, AI/NA = American Indian/Native American

Table 3 summarizes the statistically significant pairwise tests of ACE component score prevalence ratios. For each of the ACE components except for emotional abuse and parental separation/divorce, at least one group pairwise comparison was significant. Differences across racial groups were most pronounced for household substance abuse, household mental illness, and parental incarceration. There were five significant prevalence ratio comparisons each for the AI/NA, Multiracial AI/NA, and Black groups, and two for Asians. White participants generally had the lowest risk of reporting ACE components. There were no significant prevalence differences between the Multiracial AI/NA group and other groups across any of the ACE components, including with the non-AI/NA Multiracial group.

**Table 3.** Summary of significant Bonferroni-corrected Tukey’s honest significance tests of ACE component prevalence ratios, Add Health 1994-2008.

| **ACE Component** | **Comparison** | **PR (95% CI)** | **p-value** |
| --- | --- | --- | --- |
| Physical abuse | Asian-White | 1.45 (1.18, 1.77) | < 0.001 |
|  | M. AI/NA-White | 1.42 (1.19, 1.69) | < 0.001 |
| Sexual abuse | Black-White | 1.64 (1.27, 2.13) | < 0.001 |
| Emotional neglect | M. AI/NA-White | 1.37 (1.18, 1.59) | < 0.001 |
| Physical neglect | Asian-White | 1.35 (1.16, 1.56) | < 0.001 |
| Mother treated violently | Black-White | 2.33 (1.47, 3.69) | < 0.001 |
| Household substance abuse | AI/NA-White | 2.59 (1.79, 3.75) | < 0.001 |
|  | M. AI/NA-White | 1.46 (1.18, 1.79) | < 0.001 |
|  | AI/NA-Black | 2.31 (1.58, 3.37) | < 0.001 |
|  | AI/NA-Asian | 5.33 (2.84, 9.99) | < 0.001 |
|  | M. AI/NA-Asian | 3.00 (1.69, 5.31) | < 0.001 |
| Household mental illness | Black-White | 2.03 (1.71, 2.41) | < 0.001 |
|  | AI/NA-White | 3.65 (2.22, 6.02) | < 0.001 |
|  | AI/NA-Asian | 3.96 (2.10, 7.48) | < 0.001 |
| Parental incarceration | Black-White | 1.83 (1.56, 2.15) | < 0.001 |
|  | M. AI/NA-White | 1.54 (1.23, 1.93) | < 0.001 |
|  | Black-Asian | 4.54 (1.96, 11.1) | < 0.001 |

Abbreviations: PR = Prevalence Ratio M. = Multiracial; AI/NA = American Indian/Native American

## Discussion

Our study tested the hypothesis that the mean ACE score and prevalence of ACE score components is significantly higher among Multiracial participants with AI/NA ancestry than those without AI/NA ancestry. This hypothesis was definitively not supported by the summary ACE score data, as there were no significant differences between either the mean score or prevalence of components between the two Multiracial groups.

The lack of support for this hypothesis suggests that the elevated mean ACE score among Multiracial people may be due to something other than the legacy of structurally racist conditions, and that there may be experiences unique to the Multiracial population not reflected in our data. For example, studies show that in addition to traditional forms of racism, Multiracial people also experience *monoracism*, a system of racial subordination that privileges single-race identification.^43,44^ Monoracism can be perpetuated by strangers, but evidence suggests that monoracism is also a feature of many interracial and Multiracial family structures.^45–47^ It is plausible that certain forms of ACEs could be a result of monoracism, particularly forms of abuse or neglect. Another potential explanation is implicit and explicit racism that interracial couples cite as a source of relationship stress, which some have hypothesized to lead to elevated rates of intimate partner violence, divorce, and household dysfunction among interracial families.^48–52^ However, it is important to acknowledge that the source of such social stressors are exogenous to interracial couples and not an inherent feature of interracial relationships. Monoracism has received limited attention in public health literature, but could help explain disparities in health between monoracial and Multiracial people.

This study had limitations. First, ACEs are defined as events occurring before the age of 18, but several ACEs domains can only be constructed using Add Health data with questions only asked during Wave 1 when participants were ages 12-19. Therefore, observations for those ACE domains were censored for any participants that turned 18 after Wave 1. This is particularly problematic for the parental separation or divorce ACE, given how common divorce is in the US population. ACE scores may thus be biased downwards, especially for younger participants that had less under-18 observation time in Wave 1. Second, small sample sizes, especially for the AI/NA population, resulted in wide confidence intervals, making interpretation of study results challenging. Repeating this analysis in a larger dataset could reduce some of the random error due to Add Health’s relatively small study size. Third, if there is indeed an association between mean ACE score and race, the removal of the “Other” race option during Wave 3 of Add Health may have unintentionally introduced differential misclassification that could have affected results in unpredictable ways, particularly for Multiracial people who had previously endorsed the “Other” racial group. Finally, given that ACE scores of 4 or greater are associated with increased risk across the greatest number of outcomes, an alternative approach to this study could be to dichotomize the ACE score into “low” (0-3) and “high” (4+) categories.

This study is the first to demonstrate that Multiracial people with and without AI/NA ancestry do not have significantly different ACE scores, suggesting the high mean ACE score among Multiracial people warrants deeper exploration. Future studies should replicate this analysis and test the our hypothesis in larger datasets where participants are directly asked the ACEs questionnaire, where individual exposure to aggregate measures of structural racism are more readily available, and where race is assessed more stably over time such as in the Behavioral Risk Factor Surveillance System and the National Survey of Children’s Health. Detailed race variables should be more frequently made available in public-use versions of datasets, which could help provide insights to make significant progress in understanding Multiracial-monoracial health disparities.

## Data Availability

This analysis used restricted data (made available through a data use agreement); a public-use subset is available online.

https://addhealth.cpc.unc.edu/data/

